# Bias Amplification in Intersectional Subpopulations for Clinical Phenotyping by Large Language Models

**DOI:** 10.1101/2023.03.22.23287585

**Authors:** Ridam Pal, Hardik Garg, Shashwat Patel, Tavpritesh Sethi

## Abstract

Large Language Models (LLMs) have demonstrated remarkable performance across diverse clinical tasks. However, there is growing concern that LLMs may amplify human bias and reduce performance quality for vulnerable subpopulations. Therefore, it is critical to investigate algorithmic underdiagnosis in clinical notes, which represent a key source of information for disease diagnosis and treatment. This study examines prevalence of bias in two datasets - smoking and obesity - for clinical phenotyping. Our results demonstrate that state-of-the-art language models selectively and consistently underdiagnosed vulnerable intersectional subpopulations such as young-aged-males for smoking and middle-aged-females for obesity. Deployment of LLMs with such biases risks skewing clinicians’ decision-making which may lead to inequitable access to healthcare. These findings emphasize the need for careful evaluation of LLMs in clinical practice and highlight the potential ethical implications of deploying such systems in disease diagnosis and prognosis.

## Introduction

Artificial Intelligence (AI) based algorithms are rapidly influencing decision making in diverse domains such as autonomous vehicle navigation, fraud detection, recommender systems with healthcare being no exception. Owing to the availability of large volumes of data in the form of images, electronic signals and electronic health records (EHR), a vast number of AI models have been trained and used to influence clinicians’ decision-making process. In the present scenario, a mammoth amount of clinical notes are available containing densely rich information about a patient. However, capturing relevant information from such resources to assist clinicians in decision-making can be equally challenging. Language models have demonstrated a potential aid in addressing this challenge in a real-world setting. Language models can assist in identifying patterns in large amounts of patient data, which can help with early diagnosis and treatment of diseases. The advent of transformer architecture has caused a rampant surge in the development of sophisticated large language models (LLMs). Transformer architecture has the ability to capture vast amounts of semantic knowledge using self-attention mechanisms encoded as word embeddings. Hence, transformer-based language models achieved state-of-the-art results for a number of clinical applications [1–4]. This has led to automating tasks, improved patient outcomes and reduced economic costs in clinical settings by assisting physicians in clinical thought processes.

However, despite its promise, there is growing concern regarding the fairness of these systems because AI algorithms have been shown to generate and amplify bias in a number of settings. Identifying bias in AI algorithms related to the healthcare domain is important because it can lead to inaccurate predictions, misdiagnosis, and potentially harmful treatment recommendations for patients. Such bias can be introduced at different stages of the algorithm development and deployment process, including data collection and preparation, algorithm design, and training. If not addressed, bias can lead to disparities in healthcare outcomes and exacerbate existing inequalities in the healthcare system. In the context of healthcare, bias is defined as the systematic and unintended favoring of individuals or groups on the basis of characteristics such as gender, age, ethnicity or other factors. Biases can be introduced in clinical notes through various ways, which include recruitment or collection strategy of data, analysis of data, and misinterpretation of data by AI systems. Additionally, these biases may arise due to various other factors, such as clinicians’ implicit biases, differences in documentation practices or patient population demographics. This leads to skewed representations of the patient’s medical history or health status. The presence of such biases can affect clinical decision-making, diagnosis, treatment and outcomes. Therefore bias analysis and mitigation become even more essential in a domain as critical as healthcare because unintended bias may hinder diagnosis leading to inequitable access to healthcare services for under-represented groups. A significant amount of work has already been done in the domain of algorithmic bias. For example, African Americans have been denied loans and given longer prison sentences compared to their Caucasian counterparts. In the healthcare domain, different studies [5] have come up with different notions of bias and demonstrated the same for various modalities such as clinical notes, medical scans and electronic signals. Seyyed-Kalantari et al. [6] illustrated underdiagnosis bias during triage in the diagnosis of patients, stating that underdiagnosis is potentially worse than misdiagnosis because the patient still receives medical care in the latter case. They analyzed under-served populations for chest radiographs and further reported that intersectional groups of under-served populations, such as Hispanic Female patients, are more prone to underdiagnosis bias. Further, Zhang et. al [7] analyzed differences in the encoding of contextual embeddings for MIMIC-III dataset between marginalized and non-marginalized populations in terms of gender, ethnicity and insurance status. They showed that the majority group was always favored with regard to demographic denominators. Patient demographics such as gender, age, ethnicity and socio-economic status provide meaningful information used by clinicians. They can also lead to undesirable biases in AI predictions, which hinder access to healthcare services. Therefore, it is essential to design AI systems that capture the relevant information from demographic factors while minimizing the effect of bias.

The study of biases in intersectional groups provides insight into the complexities and interactions of social determinants of health and the health disparities across population subgroups. Notably, there has been limited literature in the field of Artificial Intelligence (AI) backed healthcare that examines intersectional bias for multiple demographic dimensions in case of clinical notes. Ogungbe et. al. [8] presented a survey of studies which illustrated the amplification of implicit bias for intersectional groups - the implicit bias of the participants was measured using IAT and the bias was shown to amplify for 2-fold (gender, age) and 3-fold demographics (gender, age, ethnicity). Another study [9] examined the bias between demographic intersections such as young men and old women. Tan et al. [10] suggested methods to evaluate intersectional biases for contextual word embeddings and showed that biases for the intersection of two demographic dimensions were greater than the individual dimensions. Lalor et al. [11] analyzed the intersectional bias in NLP related tasks across five text based datasets. It was reported that as the degree of intersection between groups increased, the Fairness Violation metric defined increased, indicating that the bias increases with degree of intersection. Given the staggering rate at which AI systems are being used by clinicians, the prevalence of biases in intersectional groups is an alarming cause of concern. The black box nature of Deep Learning models hinders explainability and affects decision making, thus preventing these intersectional groups from receiving timely and vital medical attention.

The key objectives of this study were to (i) Conduct bias analysis of LLMs across demographic dimensions such as age and gender; (ii) Study incremental bias across intersectional subgroups to aid clinicians in the decision making process. To this end, we analyzed a compendium of language models by performing clinical phenotyping on i2b2 2006 smoking and i2b2 2008 obesity datasets. Subsequently, bias analysis was conducted on segregated groups (age, gender) and intersectional subgroups (intersection of age and gender). To summarize, this study will enable clinicians and decision makers to be mindful of the implications of the biases introduced and amplified by LLMs making them more inclusive and suitable for real-world deployment.

## Methods

This study utilized two datasets for phenotyping - i2b2 2006 smoking [12] and i2b2 2008 obesity which consist of discharge summaries from Partners HealthCare. Each record was de-identified and annotated by two pulmonologists who used textual judgment and medical intuition for annotation. Disagreements were handled by obtaining judgments from two other pulmonologists and in case there was no majority vote, the record was omitted. For i2b2 2006 smoking, the records were classified into 5 categories - Past Smoker, Current Smoker, Smoker, Non-Smoker and Unknown. The obesity dataset records were classified into 15 categories focused around obesity and its comorbidities. Table-1 represents the statistics for both the cohorts along with gender and age-wise segregated numbers. Based on this strategy, the final cohort consisted of 398 training and 104 testing records for i2b2 2006 smoking. The i2b2 2008 obesity dataset consisted of 730 training and 507 testing records.

**Table 1:** Cohort statistics. Distribution of training & testing dataset across gender, age & intersection groups.

| Segregation Criterion | Criterion Value | i2b2 2006 smoking |  | i2b2 2008 obesity |  |
| --- | --- | --- | --- | --- | --- |
|  |  | Train Set | Test Set | Train Set | Test Set |
| Gender | Male | 163 (41%) | 50 (48%) | 363 (50%) | 272 (54%) |
|  | Female | 152 (38%) | 33 (32%) | 309 (42%) | 197 (39%) |
|  | NaN | 83 (21%) | 21 (20%) | 58 (8%) | 38 (7%) |
| Age Group | 0-20 | 5 (2%) | 1 (1%) | 11 (2%) | 4 (1%) |
|  | 20-40 | 46 (12%) | 12 (11%) | 35 (5%) | 22 (4%) |
|  | 40-60 | 107 (27%) | 28 (27%) | 177 (24%) | 144 (28%) |
|  | 60-80 | 134 (33%) | 40 (39%) | 299 (41%) | 192 (38%) |
|  | 80+ | 57 (14%) | 12 (11%) | 52 (7%) | 110(22%) |
|  | NaN | 49 (12%) | 11 (11%) | 156 (21%) | 35 (7%) |
| Age & Gender Intersection<br>(Young: 20-40;<br>Middle: 40-60;<br>Old: 60+ years) | Young Male | 14 (3.5%) | 5 (5%) | 8 (1%) | 4 (0.5%) |
|  | Middle Male | 32 (8%) | 8 (7.5%) | 95 (13%) | 66 (13%) |
|  | Old Male | 86 (21.5%) | 15 (14.5%) | 153 (21%) | 98 (19%) |
|  | Young Female | 24 (6%) | 5 (5%) | 28 (4%) | 17 (3%) |
|  | Middle Female | 45 (11%) | 12 (11.5%) | 87 (12%) | 92 (18%) |
|  | Old Female | 72 (18%) | 27 (26%) | 168 (23%) | 105 (21%) |
|  | Others (NaN & 0-20) | 125 (32%) | 32 (30.5%) | 191 (26%) | 125 (24.5%) |

The experimental setup has been as follows - the clinical notes were divided into chunks and each chunk was fed into a transformer based language model. A compendium of multiple BERT-based architectures trained across different corpora were fine-tuned on the i2b2 cohorts for phenotyping. The base BERT [13] model consists of 110M parameters and trained with two objectives - Masked Language Modeling (MLM) and Next Sentence Prediction (NSP). BioBERT [14] and BioClinicalBERT [15] provide domain specific BERT models trained on a large number of biomedical and clinical corpora such as PubMed articles and clinical datasets like MIMIC-III [16]. SciBERT [17] is trained on a large multi-domain corpus of scientific publications whereas UMLS-BERT [18] modifies the BERT architecture by fusing clinical semantic embeddings with the contextual embeddings. Each model outputs a contextual embedding vector based on the chunk fed to it. There were several ways to combine the chunks - adding an LSTM layer, taking element-wise maximum or taking the mean of all vectors. The mean of all contextual embedding vectors across all the chunks was shown to outperform all the methods of combining the chunks mentioned previously [19,20]. Hugging Face [21] library was used for pre-trained BERT-based language models. The models were trained for 1000 epochs for both datasets with a batch size of 64 samples per iteration. Binary Cross Entropy loss was used as the loss function and optimization was done using Adam optimizer with a learning rate of 6e-5. The BERT batch size was kept as 7, representing the number of samples one BERT model should take at a time. In this manner, each of the BERT models was finetuned on the respective cohorts and the aggregated results for each of the models was reported in Table-2.

For the purpose of bias analysis the data was firstly segregated based on age and gender of the patients. Each clinical note in both the cohorts followed a fixed structure in terms of headings - “DISCHARGE SUMMARY”, “HISTORY OF PRESENT ILLNESS” (HPI), “SECONDARY DIAGNOSIS”, etc such that the History of Present Illness(HPI) section contained information about the patients’ gender and age. Leveraging this information, a rule-based segmentation was performed on the clinical notes where first the note was divided into sections and subsequently, age and gender was extracted from the clinical notes. For gender segregation, a vocabulary of male and female-specific pronouns was constructed and searched for in the clinical note, following which the appropriate label was assigned. Ages were divided into five groups - 0-20, 20-40, 40-60, 60-80 and 80+ years as done in previous studies on bias in the healthcare domain [6]. The age is found by searching for keywords such as “year” and “age” in the clinical notes. With this segregation the cohorts were sliced on the basis of gender and age one by one and the fine-tuned language models were evaluated. The segregated cohorts were manually verified. Bias across gender and age for different models was reported and analyzed.

**Table 2:** Results on complete cohort for BERT-based language models

| Dataset | Model | Precision | Recall | F1-Score (micro) |
| --- | --- | --- | --- | --- |
| **i2b2 2006**<br>**smoking** | BERT | 0.85 ± 0.04 | 0.82 ± 0.04 | 0.84 ± 0.03 |
| | BioBERT | 0.84 ± 0.03 | 0.85 ± 0.03 | 0.84 ± 0.03 |
| | Bio-ClinicalBERT | 0.80 ± 0.04 | 0.81 ± 0.04 | 0.81 ± 0.04 |
| | SciBERT | 0.75 ± 0.04 | **0.95 ± 0.02** | 0.84 ± 0.03 |
| | UMLS BERT | **0.86 ± 0.03** | 0.87 ± 0.03 | **0.86 ± 0.03** |
| **i2b2 2008**<br>**obesity** | BERT | 0.64 ± 0.01 | 0.58 ± 0.01 | 0.61 ± 0.01 |
| | BioBERT | 0.72 ± 0.01 | 0.68 ± 0.01 | 0.70 ± 0.01 |
| | Bio-ClinicalBERT | 0.69 ± 0.01 | 0.71 ± 0.01 | 0.70 ± 0.01 |
| | SciBERT | 0.73 ± 0.01 | **0.74 ± 0.01** | **0.74 ± 0.01** |
| | UMLS BERT | **0.74 ± 0.01** | 0.70 ± 0.01 | 0.72 ± 0.01 |

Finally, population subgroups were created to analyze intersectional bias. An intersectional group refers to a population subgroup composed of a combination of two or more demographic dimensions. In the current study, two demographic dimensions - age and gender were considered. For example, all female patients above 80 years of age and all male patients under 40 years of age are examples of such intersectional groups. The metric used for bias evaluation of both multi-label classification tasks was micro F1-score because it is a balanced metric that takes into account both Precision and Recall. To compare bias between two groups belonging to a demographic, their respective micro F1-scores were compared. We propose and experimentally verify that individual demographic dimensions amplify the bias when multiple dimensions are combined together. To put it mathematically, *F1*^*g,a*^_*i,j*_ = *F1*^*g*^_*i*_ ⋂ *F1*^*a*^_*j*_ < *max*(*F1*^*g*^_*i*_, *F1*^*a*^_*j*_) where F1^g^_i_ denotes the micro F1-score for i’th gender group and F1^g^_j_ denotes the micro F1-score for j’th age group. F1^g,a^_i,j_ thus represents the micro F1-score for i’th gender group and j’th age group combined and serves as a metric to quantify intersectional bias. Therefore, if two groups that are already biased against on the basis of a single demographic dimension are combined, the resultant intersectional group will be at a greater risk of being biased against.

## Results

Based on the segregation criterion, the i2b2 2006 smoking dataset had 398 samples analyzed for biases across gender groups, whereas 442 samples were analyzed for biases across age groups. The i2b2 2008 obesity, on the other hand, had 1141 and 1046 samples analyzed for biases across gender and age groups, respectively. In the smoking dataset, 185 samples were female patients, whereas 506 samples were encountered in the obesity dataset. For the age group cohort, the 60-80 group had the highest frequency of samples - 174 and 491 across smoking and obesity datasets, respectively. For intersectional subgroups formed by combining demographic dimensions of age and gender, it was observed that young males and young females were the most underrepresented groups with counts of 19 and 29 for smoking and 12 and 45 for obesity datasets. Old males had the maximum samples (101) for smoking, whereas old females had the maximum samples (273) for obesity datasets. Middle males and middle females had numbers comparable to old males and old females for both datasets. Detailed statistics are shown in Table-1.

Profiling based on models suggested that all models achieved a micro F1-score of more than 0.81 and 0.70 for smoking and obesity datasets respectively. The performance of models decreased on the obesity dataset owing to the fact that the obesity dataset consisted of 15 output classes concerning obesity and associated ailments, whereas smoking consisted of only 5 output classes. Therefore, clinicians need to be more vigilant while assessing decisions made by these models on obesity-related cohorts. Table-2 demonstrates the metric evaluation of different models on phenotyping tasks. UMLS-BERT was used to perform further analysis on bias and intersectional subgroups because it was the best-performing model for both datasets and has the highest likelihood to be deployed in a practical setting to assist a clinician’s decision-making process.

Bias analysis on cohorts segregated by a single demographic dimension for age and gender were illustrated in Figure-1. For gender-based segregation, i2b2 2006 smoking dataset had ⅘ models which were biased in favor of male patients compared to female patients with a mean difference of nearly 6% in micro F1 value. The i2b2 2008 obesity dataset had all models biased in favor of male patients compared to female patients with a mean difference of nearly 3.4% in micro F1 value. For age-based segregation, patients belonging to age groups 20-60 and 60-80 were compared as both groups had a similar number of training records. The i2b2 2006 smoking dataset had ⅘ models biased in favor of patients aged 20-60 years as compared to the patients aged 60-80 with a mean difference of nearly 4.8% in micro F1 value. In the case of the i2b2 2008 obesity dataset, a complete reversal of trend was observed with ⅘ models being biased against patients aged 20-60 years as compared to the patients aged 60-80 with a mean difference of nearly 4.4% in micro F1 value.

Table-3 demonstrates gender and age-wise segregated results for both the datasets for selected groups. Table-4 shows the results for intersectional groups formed by groups depicted in Table-3. The intersectional groups were created for each dataset by combining demographic dimensions of gender and age to form four categories for each dataset - middle male, young female, middle female and old female for i2b2 2006 smoking and young male, old male, young female and middle female for i2b2 2008 obesity where ‘young’ implies ages between 20-40, ‘middle’ implies ages between 40-60 and ‘old’ implies ages >60 years. These categories were chosen because they are the most susceptible to smoking and obesity-related ailments [22–26]. It was observed that for i2b2 2006 smoking dataset ¾ intersectional subgroups - middle male, young female and old female - exhibited an amplification in bias compared to individual demographic dimensions. On the contrary, the fourth intersectional subgroup (middle females) exhibited a reduction in bias compared to individual demographic dimensions. Similar trends were observed for i2b2 2008 obesity dataset where ¾ intersectional subgroups - young male, young female and middle female - exhibited an amplification in bias. In contrast, the fourth intersectional subgroup (old male) exhibited a trend reversal owing to a reduction in bias.

**Table 3:** Segregated Results for smoking (left) & obesity (right) datasets via UMLS BERT

| Demographic Dimension | Subgroup Name | Micro F1-Score | Demographic Dimension | Subgroup Name | Micro F1-Score |
| --- | --- | --- | --- | --- | --- |
| **Age** | Young Aged | $0.88 \pm 0.08$ | **Age** | Young Aged | $0.67 \pm 0.04$ |
| | Middle Aged | $0.92 \pm 0.04$ | | Middle Aged | $0.7 \pm 0.02$ |
| | Old Aged | $0.83 \pm 0.05$ | | Old Aged | $0.74 \pm 0.02$ |
| **Gender** | Male | $0.87 \pm 0.04$ | **Gender** | Male | $0.75 \pm 0.01$ |
| | Female | $0.81 \pm 0.05$ | | Female | $0.7 \pm 0.01$ |

**Table 4:** Bias Amplification for Intersectional Groups via UMLS BERT Dataset Category Name Intersectional Results

| Dataset | Category Name | Intersectional Results |
| --- | --- | --- |
| **i2b2 2006 smoking** | Middle Male | $0.76 \pm 0.13$ |
| | Young Female | $0.8 \pm 0.18$ |
| | Middle Female | $0.92 \pm 0.08$ |
| | Old Female | $0.75 \pm 0.07$ |
| **i2b2 2008 obesity** | Young Male | $0.54 \pm 0.13$ |
| | Old Male | $0.76 \pm 0.02$ |
| | Young Female | $0.67 \pm 0.04$ |
| | Middle Female | $0.69 \pm 0.02$ |

**Figure 1.**
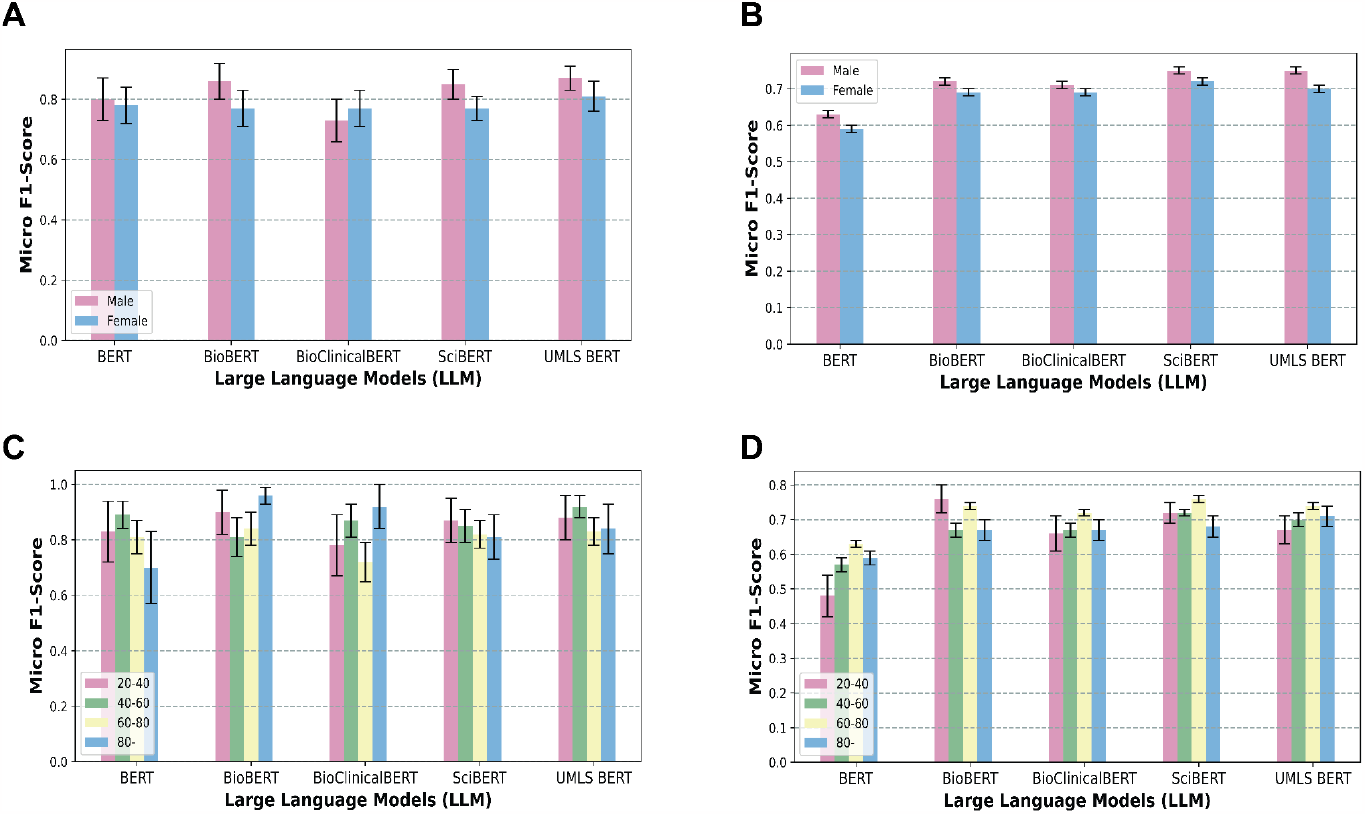
Bias Analysis across Age and Gender for Smoking & Obesity datasets. Micro F1-score of language models on Smoking and Obesity datasets w.r.t (A,B) Gender Distribution and (C,D) Age Distribution

## Discussion & Conclusion

Applications of AI systems in clinical settings extend from critical care to triage leading to reduced clinical fatigue [27]. However, the systematic unfavouring of certain population subgroups by AI systems is an alarming cause of concern because it delays and/or denies equitable access to healthcare facilities. In this work, bias analysis is conducted on clinical phenotyping tasks across two datasets - smoking and obesity. The study’s primary objective is to highlight the role of language models in the systematic amplification of bias in clinical diagnosis between diverse population subgroups. Prior work on age and gender-based segregation for clinical notes [7] and images [6] highlighted that the overrepresented (majority) group always had better metrics than the underrepresented group (minority). However, in this study, it was observed that despite having an identical number of training records, the models exhibited bias between subgroups. For gender-based segregation in both datasets, the models are always biased in favor of male patients compared to female patients despite the training cohorts having similar counts of both population subgroups. Therefore, for smoking and obesity-related comorbidities, clinicians need to be more observant for female patients irrespective of the frequency of historical data. Similarly, a significant demarcation was observed while assessing models across different age groups. The 20-60 age group is more susceptible to smoking [25,26], and the high efficacy of the models across this group enables clinicians to make better decisions. However, a dent in the performance for the 60-80 age group further validates the idea that clinicians need to be more vigilant while prognosticating such vulnerable groups. Notably, for the obesity dataset, ⅘ models showed the best efficacy for the 60-80 age group. Contrastingly, the models performed poorly for the age group >80 years. This age group is highly susceptible to obesity and also vulnerable because suffering from obesity in this age group might imply suffering from its associated comorbidities [22].

Intersectional bias was analyzed by creating population subgroups using age and gender as the demographic dimensions. Prior literature [6] highlighted that the intersection of two underrepresented groups had a higher underdiagnosis rate than the individual groups for image modality (chest radiographs). Subsequently, a study demonstrated an increase in bias as the number of demographic dimensions increased for textual dataset [11]. Our findings are very concerning from a clinical perspective as in majority cases intersectional subgroups exhibit an amplification in bias. It suggested that for the smoking dataset, the most vulnerable group - middle-aged males [25,26] had shown a higher bias compared to only middle-aged patients or only male patients. Notably for the obesity dataset, the most vulnerable group - middle-aged females [23,24,28] showed a minor increase in bias. Whereas young males, though less vulnerable than females, showed a drastic increase in bias in comparison to only young patients or only male patients.Yet, there are certain subgroups which exhibited a trend reversal (reduction in bias). Therefore, the inference of this study is crucial for clinicians leveraging these state-of-the-art language models in clinical decision-making. Clinicians should be aware of the pitfalls of these language models and sensitive towards subgroups that may be clinically misrepresented. Thus, they can make informed decisions and ensure fairness in the treatment of incoming patients. In prior studies on chest radiographs [6] and non-clinical text [11], underrepresented population subgroups were the ones being underdiagnosed by AI systems. However, in our study, the population subgroups that exhibited an amplification in bias by language models were not always underrepresented in the training cohorts.

One key limitation of this study is that we have demonstrated the fairness of language models based on a single clinical outcome. For broader applicability and a robust implementation of models, bias analysis needs to be performed across multiple tasks. Hence, in future work, we would like to extend this analysis to other clinical prognosticating outcomes such as ICU Readmission, ICU Length of Stay and ICU Mortality Prediction. The findings of this study elucidate the need for proactive measures to identify and mitigate biases in Language Models used for clinical decision-making. As shown, intersectional biases can be amplified by state-of-the-art language models when applied to clinical notes for phenotyping. This can lead to potential inequities in healthcare access and outcomes for certain population subgroups. It highlights the need for continued research and development to ensure that language models are not exacerbating existing biases in healthcare. Recognizing and addressing these biases is essential to ensure equitable healthcare for all individuals, regardless of their demographic characteristics. This asserts the development of bias-aware and explainable language models. Intervention measures can include (i) improving data collection and annotation processes (ii) incorporating diverse perspectives of various stakeholders in the development of language models (iii) augmenting training data to ensure representativeness of all population groups (iv) develop algorithms to explicitly account for intersectionality of demographic dimensions (v) leverage interpretability tools to address biases in real-time. The overarching message of this study is to use language models as a tool to augment clinicians’ decision-making process and improve patient outcomes. Furthermore, this should be done in a manner that minimizes clinical burden, reduces the potential for bias and ensures fairness for all individuals.

## Data Availability

All data produced in the present study are available upon reasonable request to the authors

## Acknowledgements

Tavpritesh Sethi acknowledges support from the DBT Project BT/PR/34245/AI/133/9/2019.

